# SARS-CoV-2 Serosurveillance Reveals Pre-pandemic Cross-Reactivity and Pandemic Seroprevalence Trends in Senegal

**DOI:** 10.1101/2025.10.20.25337295

**Authors:** Mouhamad Sy, Ian Baudi, Ibrahima M. Ndiaye, Mariama Toure, Amy Gaye, Tolla Ndiaye, Aida S. Badiane, Awa B. Deme, Jules Gomis, Daba Zoumarou, Mouhamadou Ndiaye, Khadim Diongue, Mame Cheikh Seck, Djiby Sow, Ngayo Sy, Mouhamadou A. Diallo, Marietou F. Paye, Pardis C. Sabeti, Katherine J. Siddle, Daouda Ndiaye

## Abstract

The relatively mild impact of COVID-19 in sub-Saharan Africa has raised questions about the role of pre-existing immunity in the region. One hypothesis for this unexpected observation is the presence of pre-existing cross-protective immunity, potentially induced by prior exposure to seasonal and zoonotic coronaviruses. However, the prevalence and functional relevance of such antibodies in the Senegalese population are not fully known.

To investigate this, we conducted a cross-sectional seroprevalence study using 822 plasma samples collected in Senegal before (2017–2019) and during (2020–2022) the pandemic, across regions of high (Kédougou) and low (Thiès) malaria endemicity. Samples were screened for anti-SARS-CoV-2 spike 1 IgG using enzyme-linked immunosorbent assay (ELISA), and a subset of the pre-pandemic IgG-positive samples was further tested for neutralizing activity using a surrogate virus neutralization test (sVNT).

Pre-pandemic SARS-CoV-2 IgG positivity was 39.1% [34.6 - 43.7]. No significant differences were observed in terms of age, sex, region, or malaria status. However, only 5.1% of pre-pandemic IgG-positive samples showed neutralizing activity, with 1.3% [0.1 - 6.7] in Kédougou and 9.2% [4.5 - 17.8] in Thiès. During the pandemic, IgG seroprevalence increased from the baseline around 40% in 2020 (37.3 % [27.7 - 48.1] in Kedougou and 50%[29.03 - 70.9%] in Thies), peaking near 99% of the study population by 2022 with 98.2% [93.8 - 99.5] in Kedougou and 98.8% [93.6 - 99.7] in Thies.

These results indicate widespread pre-pandemic cross-reactivity to SARS-CoV-2 in Senegal, likely driven by exposure to related coronaviruses. However, their poor neutralizing activity implies limited cross-protection. These findings highlight the need for further investigation into the origins, nature, and immunological significance of these cross-reactive antibody responses.

## Background

Since it emerged in 2019, there have been nearly 800 million reported cases of Coronavirus Disease 19 (COVID-19) (∼10% of the global population) and over 7 million deaths worldwide.^1^ The African region was predicted to experience higher COVID-19 case loads due to weaker health infrastructures and other competing public health priorities (like malaria, TB, and HIV)^2^. However, the region has instead reported a disproportionately low number of cases and overall reduced severity^2^. African countries reported significantly lower case rates; roughly 10 million COVID-19 cases, representing just 0.7% of the continent’s population^1^, in contrast to 281 million cases in Europe, 193 million cases in America, and 55 million in Southeast Asia^1^.

In Senegal, between March 2020 and May 2025, 90,000 cases (0.5% of the Senegalese population) and just under 2000 deaths had been reported^3^. This number likely reflects an underestimation of the true disease burden due to the limited availability of testing in much of the country. Indeed, a nationwide survey conducted in October–November 2020 reported a seroprevalence of 28.4%^4^. It remains unclear whether the relatively lower number of confirmed infections in Senegal is solely attributable to limited testing^5^ or reflects a reduced susceptibility to infection or a lower severity of disease^6^.

Several hypotheses have been proposed to explain the comparatively mild impact of COVID-19 in sub-Saharan Africa, including pre-existing cross-reactive immunity from exposure to other human coronaviruses (hCoVs) such as 229E, NL63, OC43, HKU1, or previous SARS-CoV infection^7–9^. SARS-CoV-2 shares approximately 79% nucleotide sequence identity with the original SARS-CoV^10^, and several studies, including samples from African countries and other localities, have reported evidence of cross-reactive antibodies to SARS-CoV-2 in pre-pandemic samples^7,9,11^. Non-coronavirus infections, including malaria, have also been suggested to cross-reactivity^12^. However, the extent to which these cross-reactive antibodies may offer immunological protection has been less thoroughly investigated^13^.

Understanding the prevalence and functional capacity of such antibodies in the Senegalese populations is critical to interpreting the country’s COVID-19 trajectory. In this context, we conducted a cross-sectional seroprevalence analysis using plasma samples collected in Senegal before the pandemic (2017–2019) and during the pandemic period (2020–2022). Our primary objective was to estimate the prevalence of IgG antibodies targeting SARS-CoV-2 in both timeframes. Lastly, we evaluated the neutralizing capacity of the detected antibodies using a surrogate virus neutralization test (sVNT), to assess whether seropositive samples, particularly those from the pre-pandemic period, contained functional antibodies capable of inhibiting SARS-CoV-2.

## Methods

### Study Population and Ethics Statement

Samples were collected from clinics as part of malaria and non-malaria surveillance in Kedougou, in south-eastern, and Thies, in western, Senegal. This collection was coordinated under the Malaria Genomic Surveillance Study, the West African Research Network for Infectious Diseases (WARNID), and the Human Heredity & Health in Africa (H3A) initiative projects. WARNID and H3A samples were collected from individuals with and without malaria. All samples from the Malaria Genomic Surveillance Study were malaria positive. Malaria cases were defined as those with symptomatic, uncomplicated *Plasmodium falciparum* infections detected by rapid diagnostic tests (RDTs) and microscopy. The study was approved by the National Ethics Committee of the Ministry of Health in Senegal (Protocol SEN 14/49). All samples were collected with written informed consent, in accordance with the ethical guidelines of both institutions. Blood specimens were collected between 2017 and 2022 through passive case detection from patients aged 6 months or older. Study staff obtained informed consent from all participants, or a parent/legal guardian for those under 18 years of age, along with assent from the minor.

A total of 822 plasma samples were identified for this study: 301 from Thiès and 521 from Kédougou. Pre-pandemic samples (n = 437) were collected between 2017 and 2019, and pandemic samples (n = 385) were those collected between July and November during each year from 2020 to 2022.

### Laboratory analysis

#### Human Anti SARS-CoV-2 Virus Spike 1 IgG ELISA

To assess exposure to SARS-CoV-2 or cross-reactivity to the virus’s antigen, we used an enzyme-linked immunosorbent assay (ELISA) to screen both pre-pandemic and pandemic samples for IgG antibodies targeting the SARS-CoV-2 Spike 1 protein. The Alpha Diagnostic International ELISA kit (Catalog #RV-405200) was used according to the manufacturer’s instructions. Plasma samples were diluted 1:500, experimentally validated within the recommended ratio range to optimize antibody detection. Optical density (OD) result values were interpreted as positive or negative based on the threshold index method using the 1U/mL calibrator. The index was calculated by dividing the sample’s background-corrected OD by the corrected calibrator OD.

### SARS-CoV-2 Surrogate Virus Neutralization Test (sVNT)

To assess the functional capacity of the antibodies among samples that tested positive for anti-SARS-CoV-2 IgG, we employed the GenScript SARS-CoV-2 Surrogate Virus Neutralization Test (sVNT) Kit according to the manufacturer’s instructions, using a 96-well microplate format. The sVNT assay is species and isotype-independent, making it applicable to a wide range of samples, including human plasma. The assay detects neutralizing antibodies that block the interaction between the receptor-binding domain (RBD) of the SARS-CoV-2 spike protein and the angiotensin-converting enzyme 2 (ACE2) receptor on host cells. After incubating the plasma and serum samples with an RBD-ACE2 recombinant complex, neutralizing antibodies will inhibit the binding of the RBD to the ACE2 receptor. The inhibition rate is calculated according to the following formula:

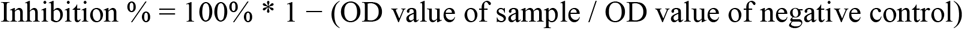

Positive results for inhibition by SARS-CoV-2 neutralizing antibodies were defined by OD values greater than or equal to 30%. Results below 30% were considered negative for neutralizing antibodies. The sVNT was performed in duplicate to ensure accuracy.

Controls for the test included 8 IgG-positive samples from the pandemic period (n = 4 from Thiès and n = 4 from Kédougou), along with 10 IgG-negative samples (7 pre-pandemic and 3 pandemic). Furthermore, nine pre-pandemic samples from the USA were used as negative controls to assess SARS-CoV-2 spike neutralization in pre-pandemic IgG-positive samples.

### Statistical Analysis

Laboratory data, including absorbance values, age, gender, and malaria status, were recorded using Microsoft Excel (Microsoft, Redmond, WA, USA). Statistical analyses and data visualization were generated using Python packages, including pandas, Matplotlib, and seaborn (https://seaborn.pydata.org), within Jupyter notebooks. Seroprevalence proportion differences between gender, locality, and period (pre-pandemic and pandemic) were assessed using Fisher’s exact test. The seroprevalence difference between age groups was assessed using the t-test. The 95% Confidence Interval was estimated using the Wilson Score. The significance threshold was set at p < 0.05.

## Results

### Demographic Characteristics

This study included a pre-pandemic cohort of 437 participants and a pandemic cohort of 385 participants, with females representing 33.6% and 39.5% of each cohort, respectively (Table 1). Overall, the participants’ ages ranged from 2 to 73 years. The median ages of the cohorts were 18 years (IQR 12-28) for the pre-pandemic and 20 years (IQR 13-31) for the pandemic cohort (Table 1). Both groups consisted of younger individuals, with over half of the participants in each cohort being 20 years of age or younger, i.e., 58.3% pre-pandemic and 52.5% during the pandemic.

**Table 1:** Study population characteristics.

|  | Thies (n = 301) | Kedougou (n = 521) | Total (N = 822) |
| --- | --- | --- | --- |
| **Pre-pandemic** |  |  |  |
| Sample collection Year | 2017 - 2019 | 2017 - 2019 |  |
| Number of samples (%) | 198 (65.8) | 239 (45.9) | 437 (53.2) |
| Age Median (IQR) | 22 (13-34) | 15 (11-23) |  |
| Age group, n(%) |  |  |  |
| ≤ 20 years | 86 (43.4) | 169 (70.7) | 255 (58.3) |
| 21 - 40 years | 83 (42) | 62 (26) | 145 (33.2) |
| > 40 years | 29 (14.6) | 8 (3.3) | 37 (8.5) |
| Female, n (%) | 44 (22.2) | 103 (43.1) | 147 (33.6) |
| Unreported sex |  | 1 (0.4) | 1 (0.2) |
| Malaria Positive, n (%) | [2] 154 (77.8) | 239 (100) | 393 (89.9) |
| Anti-SC2 IgG pos (%) (95% IC) | 40.9 [34.3 – 47.8] | 37.7 [31.8 - 43.9] | 39.1 [34.7 - 43.8] |
| **Pandemic** |  |  |  |
| Sample collection Year | 2020 & 2022 | 2020 - 2022 |  |
| Number of samples (%) | 103 (34.2) | 282 (54.1) | 385 (46.8) |
| Age Median (IQR) | 20 (15-33) | 20 (13-30) |  |
| Age group, n (%) |  |  |  |
| ≤ 20 years | 55 (53.4) | 147 (52.1) | 202 (52.5) |
| 21 - 40 years | 32 (31.1) | 111 (39.4) | 143 (37.1) |
| > 40 years | 16 (15.5) | 24 (8.5) | 40 (10.4) |
| Female, n (%) | 19 (18.4) | 133 (47.1) | 152 (39.5) |
| Malaria Positive, n (%) | 103 (100) | 282 (100) | 385 (100) |
| Anti-SC2 IgG pos (%) (% 95% IC) | 80.6 [71.9 - 87.1.6] | 72.3.8 [66.9 - 77.2] | 74.7 [70.2 - 78.8] |

### Seroprevalence and cross-reactive anti-SARS-CoV-2 IgG

Given the endemicity of malaria in Senegal, we first assessed whether acute malaria infection influenced the performance of the SARS-CoV-2 IgG ELISA. Previous reports have suggested possible cross-reactivity in malaria-positive individuals^12,14^. Only the 2018 Thies data included both malaria-positive (n = 46) and negative samples (n = 37). The seroprevalence was 50.0% (95% CI : 36.1 – 63.9) in malaria-negative individuals and 54.1% (95% CI : 38.4 – 69.0) in malaria-positive individuals, showing no statistically significant association (p = 0.8258) (Figure 1A). These results suggest that the current malaria status did not significantly affect assay specificity in this study.

**Figure 1.**
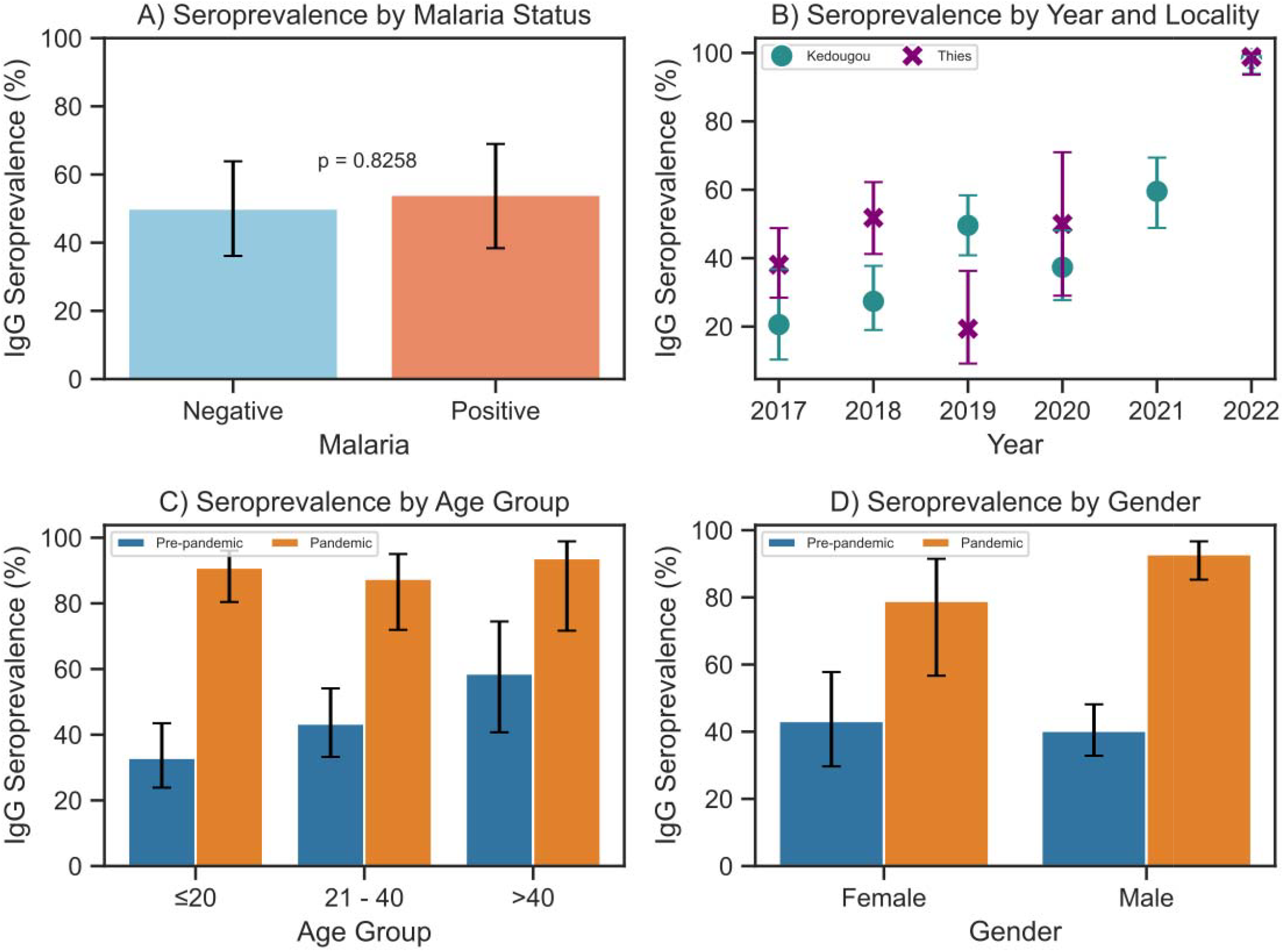
Anti–SARS-CoV-2 IgG prevalence in pre-pandemic and pandemic periods. A) shows the seroprevalence of SARS-CoV-2 according to malaria status in Thies (samples collected in 2018). B) represents the Anti–SARS-CoV-2 IgG prevalence in Thies and Kedougou by year. C) IgG seroprevalence by age group and gender between pre-pandemic (2017–2019, blue) and D) by (2020– 2022, orange) periods. All figures display error bars of 95% confidence interval and the Fisher’s exact test p-value.

Of the 437 pre-pandemic samples tested, the overall SARS-CoV-2 IgG positivity was 39.1 % [95% CI: 34.7 – 43.8], with a slightly higher seroprevalence in Thies (40.9% [95% CI: 34.3 – 47.9]) compared to Kedougou (37.7% [95% CI: 31.8 – 43.9], Table 1). As shown in Figure 1B, the proportion of participants testing positive for SARS-CoV-2 cross-reactive antibodies trended up in Kedougou between 2017 and 2019 cohorts from 20.6% to 49.6%, while in Thies it initially went up through 2018, then went down in 2019 (38.1% to 51.8% then down to 19.4%. Age-stratified seroprevalence was 38.8% [95% CI: 33.1 – 44.9] among individuals aged ≤ 20 years, 36.6% [95% CI: 29.2 – 44.6] among those aged between 21 and 40 years, and 54.1% [95% CI: 38.4 – 69.0] in participants over 40 years (supplementary fig 2). No association was found between cross-reactive anti-SARS-CoV-2 IgG positivity and locality, gender, or across age subgroups. During the pandemic, overall SARS-CoV-2 IgG seroprevalence increased markedly to 74.7% (95% CI: 70.2 – 78.8), which was significantly higher than the pre-pandemic level of 39.1%. Regional prevalence remained slightly higher in Thies (80.6% [95% CI: 71.9 – 87.1]) compared to Kedougou (72.3% [95% CI: 66.9 – 77.2]) (Table 1), though not significantly. As shown in Figure 1B, the seroprevalence increased from approximately 40% in 2020 to nearly 99% of the study population by 2022. This trend likely reflects cumulative exposure due to natural infection and/or vaccination during the pandemic. Also to note, seroprevalence during the early pandemic years 2020 closely mirrored baseline pre-pandemic level before increasing significantly and peaking in 2022. Seroprevalence in males (78.1% [95% CI : 72.4 – 82.9]) was slightly higher than in females (69.1% [95% CI : 61.3 – 75.9%]), though this difference was not statistically significant. Similarly, age-stratified seroprevalence did not differ significantly across subgroups, i.e., among ≤ 20 year olds (73.3% [95% CI : 66.8 – 79.2]), 21-40 year olds (76.2% [95% CI : 69.2 – 83.2]), and the over 40 year olds (75% [95% CI : 60.0 – 87.5]).

**Figure 2.**
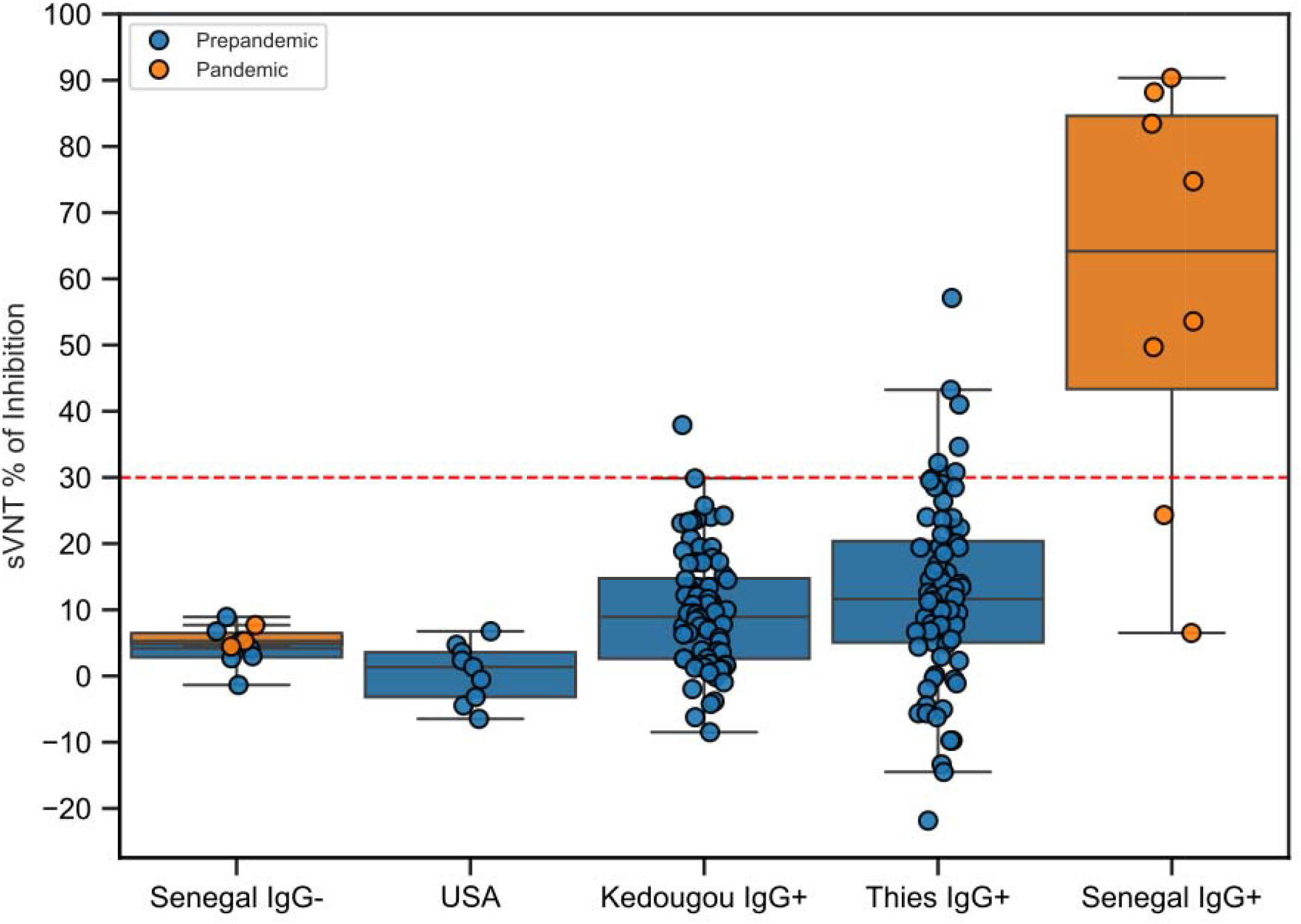
SARS-CoV-2 sVNT inhibition percentage by group and period. Each dot represents an individual tested sample. The horizontal dashed red line marks the cutoff at 30% inhibition. Boxplots show the distribution of inhibition values within each group, with overlayed points representing individual samples. Groups include pre-pandemic localities (USA, Kedougou IgG+, Thies IgG+), pandemic samples stratified as Senegal *IgG+*, and ELISA-negative controls (Senegal *IgG-*).

### Evaluation of neutralizing antibody activity in cross-reactive IgG-positive samples

Out of the 170 SARS-CoV-2 IgG□positive pre-pandemic samples, 156 samples (n = 80 from Thiès and n = 76 from Kédougou) were selected randomly and tested for antibody neutralization activity by a commercial surrogate viral neutralization assay. For controls, we included 8 IgG-positive samples from the pandemic period, 10 IgG-negative mixed samples from pre-pandemic and pandemic periods in Senegal, and 9 pre-pandemic samples from the USA. Among the pandemic-period controls, 6 of 8 neutralized the SARS-CoV-2 spike at a cut-off of > 30% inhibition used in this study. No neutralization was observed in the negative control samples from Senegal or those from the USA (Figure 2).

Of the 156 samples tested, Figure 2 shows only 8 (5.1%) were classified as positive for neutralizing antibodies. Thies showed a marginally higher prevalence of neutralizing antibodies at 9.2% [95% CI: 4.5 – 17.8] compared to Kedougou at 1.3% [95% CI: 0.1 – 6.7]. No significant associations were observed between neutralizing antibody status and age or sex. By age group, prevalence was highest among individuals aged 21– 40 years at 9.6% [95% CI: 4.2 – 20.6], followed by those over 40 years at 5.0% [95% CI: 0.3 – 23.6], and those under 20 years at 2.4% [95% CI: 0.4 – 8.3]. By sex, males had a neutralizing antibody prevalence of 5.8% [95% CI: 2.7 – 12.1], while females had a prevalence of 3.8% [95% CI: 0.7 – 12.8] (Figure 2, Table 2).

**Table 2:** Prevalence of Inhibition Pre-pandemic.

|  | Total (156) | Prevalence n (%) | CI Lower | CI Upper |
| --- | --- | --- | --- | --- |
| **Locality** |  |  |  |  |
| Thies | 76 | 7 (9.2) | 4.5 | 17.8 |
| Kedougou | 80 | 1 (1.3) | 0.1 | 6.7 |
| **Age Group** |  |  |  |  |
| <20 | 84 | 2 (2.4) | 0.4 | 8.3 |
| 20-40 | 52 | 5 (9.6) | 4.2 | 20.6 |
| >40 | 20 | 1 (5.0) | 0.3 | 23.6 |
| **Gender** |  |  |  |  |
| Female | 53 | 2 (3.8) | 0.7 | 12.8 |
| Male | 103 | 6 (5.8) | 2.7 | 12.1 |

## Discussion

This study provides robust evidence of pre-pandemic seropositivity to SARS-CoV-2 spike protein in Senegal, indicating that a portion of the population had prior exposure to antigenically related viruses. Among the samples collected between 2017 and 2019, the seroprevalence of cross-reactive IgG antibodies was 39.1%, with comparable levels between the regions of Thiès (40.9% [95% CI : 34.3 – 47.9]) and Kédougou (37.7% [95% CI : 31.8 – 44.0]). However, neutralization testing revealed that only a small fraction (5.1%) of these pre-pandemic IgG-positive samples exhibited neutralizing activity against SARS-CoV-2, indicating limited functional cross-protective potential^9,13^.

Our results are consistent with the growing evidence from sub-Saharan Africa and other regions, which reports pre-pandemic seroreactivity to SARS-CoV-2 antigens^7,11^. In Senegal, previously reported pre-pandemic IgG seroprevalence rates range widely, up to 23.5%^9^. Similar patterns have been observed in Nigeria, Kenya, Tanzania, Uganda, and Sierra Leone, where 5-52% of individuals tested positive for cross-reactive antibodies^7,11,15^. Pre-pandemic cross-reactive antibodies were more frequent in Africa (31.9%–52%)^7,8,16^ and Asia (4%–20%)^14,17^ but remained rare in Europe and the Americas (<5%). For example, only 2.4% of Canadians, 2.5% of Danish, and 3.7% of Brazilian samples were positive^13^.

Our study reinforces this evidence, while adding that neutralizing activity was uncommon, as similarly observed in female sex workers in Dakar^9^. These findings suggest that much of the observed reactivity is likely non-protective and driven by exposure to coronaviruses with shared epitopes^7,9^. The use of the spike S1 domain, a less conserved protein than others, like the nucleocapsid, in our assay supports the specificity of the observed responses. Although these antibodies were largely non-neutralizing, they may contribute to immunological memory or modulate disease severity through other immune mechanisms. In addition to a lower mortality rate (2.5%), among confirmed cases in Senegal, only 39% of COVID cases presented clinical signs^5^. This trend was also observed across much of sub-Saharan Africa. Indeed, at the beginning of the pandemic, an estimate based on WHO data indicated that the case fatality rate in Africa was very low, around 3 per 10,000 infections, compared with 13.80 per 10,000 in the Americas and 20.19 per 10,000 in Europe^18^ (H.H. Ayoub et al, 2021).

The high baseline seropositivity of around 40% observed in pre-pandemic samples poses questions about the source of the cross-reactive immune response observed. It is plausible that repeated or intermittent exposure to antigenically related but distinct coronaviruses of human or animal origin underlies these observations^19^. We and others have performed ecological surveillance that points strongly to potential zoonotic reservoirs of sarbecoviruses in bats in and around Thies. We found that about 11% of bat specimens tested positive for bat coronaviruses (batCoVs)^20^. In contrast, very few human participants screened by PCR, tested positive for betacoronaviruses, suggesting that human infection with known seasonal coronaviruses was fairly uncommon in this population. The same studies also revealed that bats were present in or near human dwellings, with around 5% of participants reporting direct contact with bats. However, further metagenomic or pan-coronavirus surveillance is required to help identify these viral sources and their immunological relevance.

Our pandemic-era results are similarly consistent with findings from other serosurveys in Senegal, Thiam *et al*., reported seroprevalence rates exceeding 40% in several regions by mid-2020, while a 2022 study in Dakar documented IgG positivity in over 90% of a university population^21^. Importantly, we observed no significant differences in pandemic-era seroprevalence by age or gender, suggesting widespread exposure and/or equitable vaccine distribution. Pre-pandemic data, however, revealed slightly higher seroprevalence in individuals over 40 years, possibly reflecting cumulative lifetime exposure to related coronaviruses^9^. This difference disappeared during the pandemic, consistent with a homogenization of exposure risk.

This study has several limitations. First, both cohorts were skewed toward younger individuals, possibly limiting conclusions about the older age groups and restricting broader generalizations. This could be explained by the fact that the samples were mostly collected as part of a study on malaria genomic surveillance. Several studies have shown that children under the age of five, as well as adolescents, tend to have a higher number of malaria cases in malaria endemic countries such as Senegal compared to adults. This is because adults generally have a more robust immune system due to past infections, which provide them with partial immunity to malaria^22,23^. Second, our serological test targeted a single antigen (spike S1), future studies could include additional targets (e.g., RBD or nucleocapsid) that may provide broader insight into cross-reactivity. For neutralization testing, only a subset of IgG-positive samples was tested using a surrogate neutralization method. Thirdly, pre-pandemic era risk factor, and pandemic era vaccination status were not systematically captured, which complicates the interpretation of the observed seroprevalence. Finally, our study focused primarily on humoral immunity and did not assess cellular immune responses. Furthermore, clinical outcomes such as disease severity or hospitalization were not evaluated. As such, it remains possible that non-neutralizing antibodies or T-cell–mediated responses contributed to protection or shaped the clinical course of infection. Future studies should incorporate PBMC analysis, protein microarrays, or VirScan-based approaches to characterize the full antibody repertoire and its relationship to exposure from other circulating pathogens, and/or map all past pathogen exposure that may have played a role in the dynamics of SARS-CoV-2 in Senegal.

In conclusion, this study highlights the presence of pre-pandemic SARS-CoV-2 cross-reactive antibodies in Senegal, shaped by regional viral ecology and possible past exposures to related coronaviruses. While most of these antibodies lacked neutralizing activity, their widespread detection suggests complex immunological interactions could be at play. Understanding the origins and functional relevance of such responses will be critical for interpreting disease patterns and guiding preparedness efforts in regions with high zoonotic and endemic disease pressures. Future work should explore broader immune mechanisms and viral reservoirs to uncover the full landscape of coronavirus exposure in Africa.

## Supporting information

Sup. Fig 1: Overall age distribution

## Contributions

M.S., I.B., M.F.P, P.C.S., K.J.S., D.N. conceived and designed the study; A.G., T.N., M.S., M.T., I.M.N., A.M.M., Y.D. acquired the data; M.S., I.B., M.S., M.T., I.M.N. analyzed the data; M.S., I.B., M.F.P., M.T., I.M.N., K.J.S. drafted and revised the manuscript with input from all authors. All authors approved the manuscript before submission.

## Data Availability Statement

The datasets generated and analyzed during the current study are included within the manuscript (and its **Supplementary Information** file). The study’s raw data can be made available from the corresponding authors upon request.

