## Supplementary material for "SARS-CoV-2 Serosurveillance Reveals Pre-pandemic Cross-Reactivity and Pandemic Seroprevalence Trends in Senegal": Sup. Fig 1: Overall age distribution

### **Supplementary or exploratory figure/table alternatives**

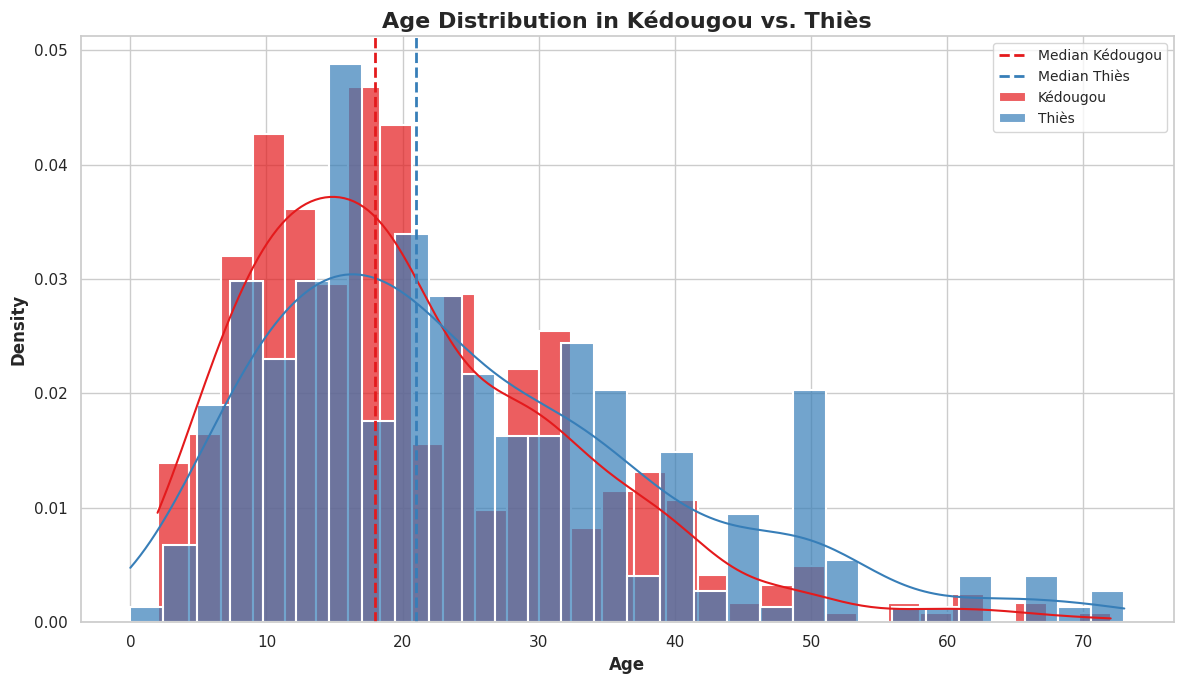

Sup. Fig 1: Overall age distribution
